# Frontal lobe fALFF measured from resting-state fMRI at baseline predicts acute treatment response in first episode psychosis

**DOI:** 10.1101/2021.07.25.21261112

**Authors:** Todd Lencz, Ashley Moyett, Miklos Argyelan, Anita D. Barber, John Cholewa, Michael L. Birnbaum, Juan A. Gallego, Majnu John, Philip R. Szeszko, Delbert G. Robinson, Anil K. Malhotra

## Abstract

While antipsychotic medications have been utilized for decades, many patients experiencing psychosis do not demonstrate a satisfactory positive symptom response, even in the first episode of illness. Resting state functional magnetic resonance imaging (rs-fMRI) has increasingly been studied as a potential biomarker of antipsychotic treatment response, but studies to date remain limited in terms of sample size and brain regions examined. The present study examined fractional amplitude of low frequency fluctuations (fALFF) derived from rs-fMRI to search the whole brain, in a hypothesis-free manner, for regions that might predict response to 12 weeks of treatments with standard antipsychotic medications (risperidone or aripiprazole). To our knowledge, the present study represents the largest first episode psychosis treatment sample studied with rs-fMRI to date (N=126). At baseline, patients who would later meet strict criteria for clinical response demonstrated significantly greater baseline fALFF in bilateral orbitofrontal cortex compared to non-responders. In a subset of 62 patients who were scanned a second time at the end of treatment, responders and non-responders demonstrated significant increases in fALFF in differing brain regions. Specifically, responders demonstrated significant increases in fALFF, from baseline to follow-up, in bilateral dorsal prefrontal cortex; prefrontal changes were not observed in non-responders or in healthy individuals (n=45) scanned at two time points. Thus, spontaneous activity in orbitofrontal cortex may serve as a prognostic biomarker of antipsychotic treatment, while spontaneous activity in the dorsal prefrontal cortex may serve as a critical target of effective medication.

---

Antipsychotic medications with activity at the dopamine D2 receptor remain the primary pharmacologic treatment for patients with psychotic disorders such as schizophrenia, despite highly variable response^1^. About one-third of patients with schizophrenia are ultimately resistant to treatment with antipsychotic medications^2^, and more than half of these lack even minimal response (20% reduction of positive symptoms) in the first episode of illness^3,4^. Moreover, a large number of patients demonstrate only partial response to antipsychotic treatment^5^, and a substantial minority of first episode patients demonstrate robust response (defined either as 50% reduction in positive symptoms^4^ or as absence of frank psychosis^6,7^). The first episode of illness is especially important clinically for at least three reasons: 1) it is the phase of illness associated with the greatest risk of suicide^8,9^; 2) it may be the best opportunity to mitigate long-term illness trajectory^10^; and 3) the onset period of late adolescence/early adulthood represents a critical period for attaining functional milestones in the transition to independence^11^. Thus, identification of individuals at risk for poor response is an important clinical goal, yet prognostic biomarkers are lacking^12^.

Furthermore, from a research perspective, identification of biomarkers for poor response to antipsychotics may point towards novel treatment targets and mechanisms not addressed by existing pharmacologic agents^12,13^. Biomarker studies conducted in first episode patients may be advantageous, compared to studies in more chronic populations, due to the ability to obtain baseline measurements unconfounded by lengthy exposure to treatment and ongoing illness processes^14^ and the availability of the full range of clinical outcomes, as compared to the abundance of partial and poor responders in samples of convenience^4^.

Resting-state functional magnetic resonance imaging (rs-fMRI) has become an increasingly utilized tool in the quest for treatment biomarkers in schizophrenia^15^. Compared to task-based fMRI, rs-fMRI is more easily performed in acutely ill patients in their first psychotic episode, and may provide information congruous to that obtained with task-based fMRI^16^. Importantly, the large majority of the energy expenditure of the brain is accounted for by spontaneous fluctuations, whereas task-related activations represent <5% of this total^17^.

Although many rs-fMRI studies of antipsychotic response have been limited to the cross-sectional comparison of treatment-responsive and non-responsive patients to each other and/or to controls^15^, Tables 1 and 2 display key parameters of all *prospective* rs-fMRI studies in first-episode psychosis to date. Table 1 includes all studies in which baseline rs-fMRI measures were examined in relation to subsequent treatment response over a period of weeks or months (we call these “baseline-biomarker” studies). Table 2 includes all studies in which patients were scanned twice, at baseline and after a period of treatment; in these studies, the *change* in rs-fMRI measures was examined in relation to change in symptoms (i.e., response) over the same time interval (we call these “change-biomarker” or “Δ/Δ” studies). While baseline-biomarker studies and change-biomarker studies are methodologically similar, they hold conceptually different roles in advancing our understanding of antipsychotic efficacy: the former hold the possibility of providing prognostic indicators for precision medicine, while the latter can identify neurocircuits that serve as targets of effective medication^18^. In general, longitudinal sample sizes for both sets of studies were small (median N=36 in Table 1, and median N=25.5 in Table 2). Notably, the two studies with relatively large sample sizes in Table 1 both included a mix of first-episode and multi-episode patients in their longitudinal studies^19,20^.

**Table 1.**
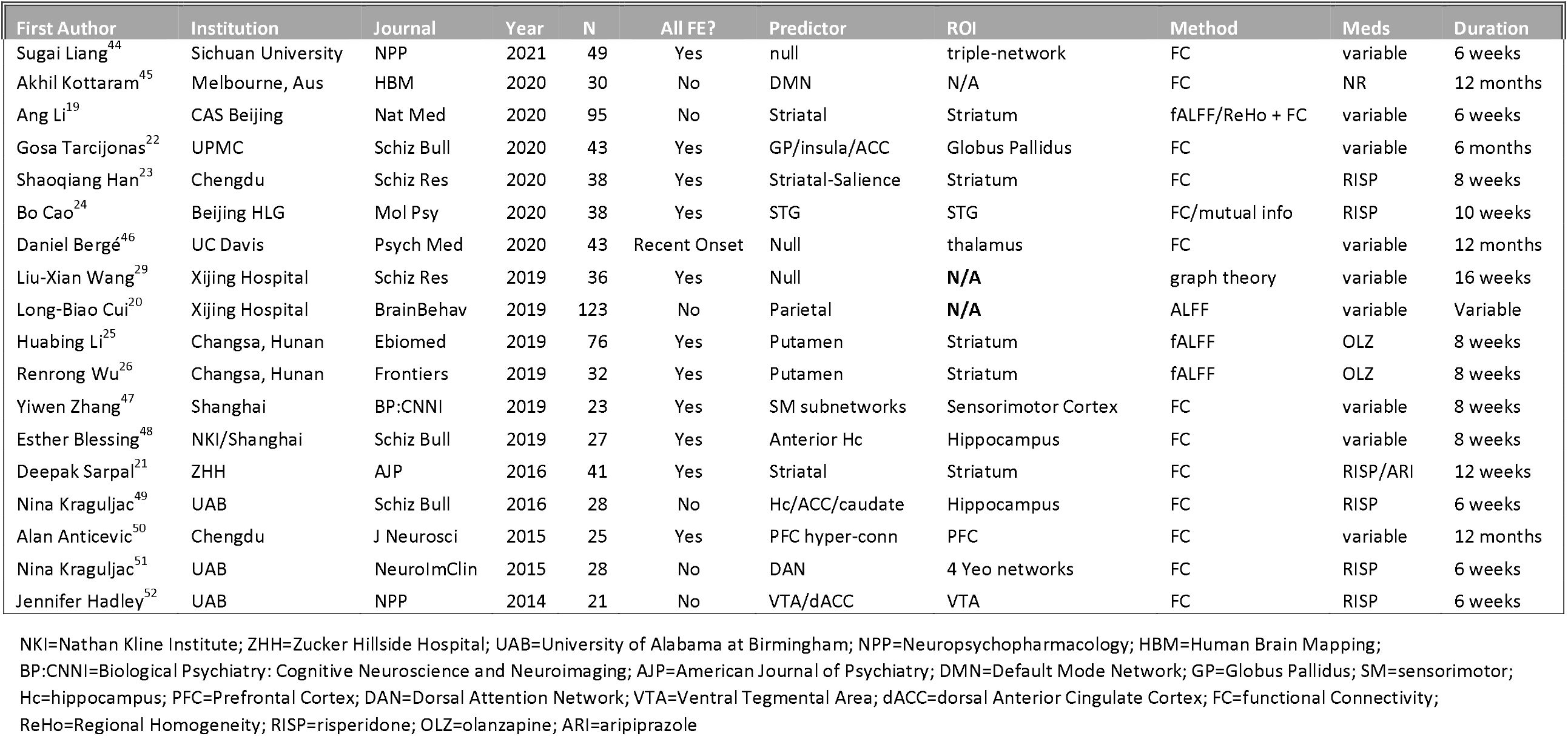

**Table 2.**
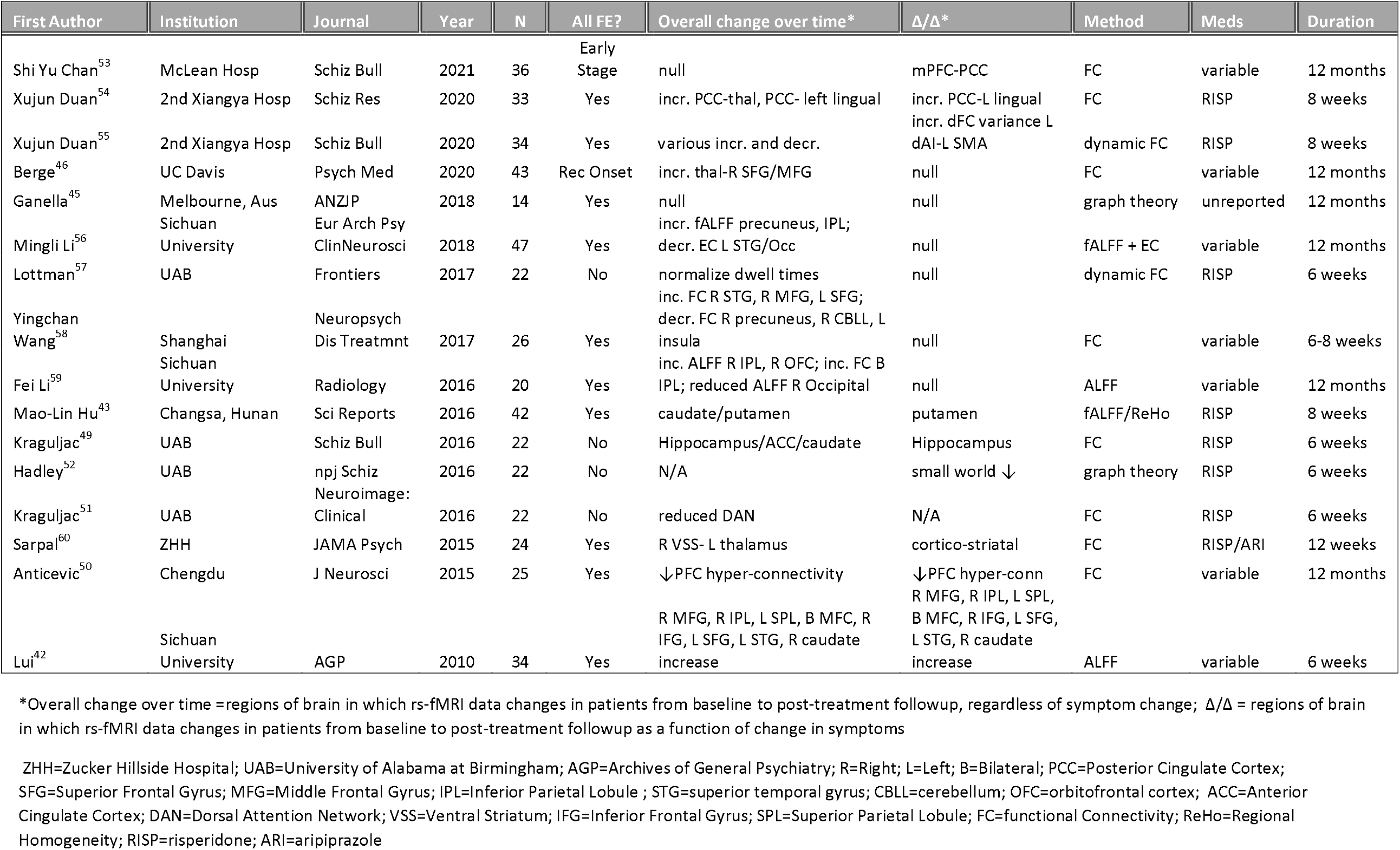

Most of the baseline-biomarker studies listed in Table 1 limit the investigation to functional connectivity of pre-defined regions of interest (ROIs). For example, we^21^ and others^19,22–26^ have focused on the corpus striatum, given the enrichment of this structure in dopamine D2 receptors, the target of all effective antipsychotics^27,28^. Only two baseline-biomarker studies, both reported from Xijing Hospital (Xi’an, China), examined the whole brain in a hypothesis-free manner. Of these, one^29^ was underpowered (n=36) and reported null results; the other^20^ included a large sub-cohort of patients with chronic illness mixed together with first-episode patients, treated with a wide range of different antipsychotic medications (including clozapine), with outcome measured after variable treatment durations.

While most of the studies in Tables 1 and 2 examined resting-state functional connectivity (FC) *across* brain regions, intrinsic resting activity *within* brain regions has been less well studied as a potential biomarker. Specifically, the fractional amplitude of low frequency fluctuations (fALFF^30^), is a measure thought to capture spontaneous neural activity, and has been shown to correlate with local brain glucose metabolism^31–34^. Importantly, regions with greater fALFF also demonstrate greater degree centrality^35,36^, a measure of connectedness derived from graph theory, suggesting that fALFF underpins (to some extent) functional connectivity of brain networks. However, relative to FC, fALFF has the advantage of being relatively less susceptible to movement artifact^37^, and fALFF has been shown to be less impacted by physiologic noise compared to an older measure of local activity, ALFF^30,38^. Moreover, fALFF has been demonstrated to be stable over the course of an fMRI session^39^, as well as reliable over time (measured in hours, weeks, or months)^38,40^, a necessary attribute for a potential biomarker^41^.

One very early study^42^ of ALFF as a change-biomarker suggested that treatment-related increases in ALFF across several cortical regions (prefrontal, dorsal parietal, and superior temporal), as well as the caudate, were correlated with symptom improvement over 6 weeks. A subsequent study using fALFF in the context of an 8-week risperidone trial provided evidence for a striatal, but not cortical, change-biomarker^43^. Two long-term (1-year follow-up) studies using ALFF or fALFF failed to find any significant change-biomarkers, although these studies did not provide data on the acute course of the initial antipsychotic trial.

The goal of the present study was to test whether measurement of fALFF can provide a baseline-biomarker and/or change-biomarker of acute (12-week) positive symptom response to antipsychotic medication in patients experiencing a first episode of psychosis. To allow hypothesis-free biomarker discovery, we utilized voxelwise measures of fALFF across the whole brain. To our knowledge, our cohort (n=126 patients with baseline and clinical follow-up, of whom 62 also had follow-up scans at 12 weeks) represents the largest prospective rs-fMRI study in first-episode psychosis to date.

## Methods

### Subjects

Subjects included 126 patients (33.3% female, mean age=22.6, SD=5.7) with first episode psychotic disorders and minimal exposure to APs (median exposure = 5 days; all patients <2 years). All subjects underwent scanning while entering 12 weeks of prospective treatment with second-generation APs (risperidone or aripiprazole). Consistent with our prior studies^6,7^, stringent treatment response criteria were applied for ratings obtained on weeks 1, 2, 3, 4, 6, 8, 10, and 12: response required 2 consecutive ratings of much or very much improved on the CGI, as well as a rating of ≤3 on psychosis-related items of the BPRS-A. By these criteria, 83 patients were classified as responders; these subjects did not differ from 43 non-responders on age, sex, scanner type (GE or Siemens), or movement during the scan (framewise displacement, FD), as displayed in Table 1. Of the 62 patients who completed a follow-up scan, 41 were responders and 21 were non-responders. In order to examine effects of time and re-scan independent of illness and treatment, we scanned 45 healthy comparison subjects (53.3% female, mean age=25.6, SD=4.5). All subjects provided written, informed consent under a protocol approved by the Institutional Review Board of the Feinstein Institutes for Medical Research at Northwell Health.

**TABLE 1.**

| **TABLE 1** | **Responders (N=83)** | **Non-Responders (N=43)** | **Statistical Test (p-value)** |
| --- | --- | --- | --- |
| **Sex**<br>**(Male/Female)** | 26/57 | 16/27 | $\chi^2(1)=0.44$ (p=0.55) |
| **Scanner Type**<br>**(GE/Siemens)** | 53/30 | 21/22 | $\chi^2(1)=2.64$ (p=0.10) |
| **Mean Age (SD)** | 22.49 (5.58) | 22.89 (5.94) | $t_{124}=0.38$ (p=0.70) |
| **Mean FD (SD)** | 0.142 (0.106) | 0.156 (0.137) | $t_{68.48}=0.58$ (p=0.56)<br>(Note: unequal variances) |

### Scan parameters

All fMRI exams were conducted on a 3T scanner (GE Signa HDx, n=77 at baseline, n=35 at follow-up); Siemens PRISMA, n=53 at baseline, n=27 at follow-up). For each patient, the follow-up scan was conducted on the same scanner as the baseline. On the Signa, the resting-state scan lasted 5 minutes, during which 150 EPI volumes were obtained (TR = 2000 ms, TE = 30 ms, matrix = 64*64, FOV = 240 mm; 40 contiguous 3mm oblique axial slices). On the PRISMA, two 7-minute 17-second resting-state runs were obtained, one each with AP and PA phase encoding directions. Resting scans contained 594 whole-brain volumes, each with 72 contiguous axial/oblique slices in the AC-PC orientation (TR=720ms, TE=33.1ms, matrix = 104×90, FOV = 208mm, voxel = 2×2×2mm, multi-band acceleration factor=8).

### Calculation of fALFF and statistical analysis

Raw resting state data were preprocessed with despiking, linear trend removal, spatial smoothing (6mm^3^ kernel FWHM), and grand mean scaling. Utilizing Fourier Transformation at every voxel, we calculated the power of BOLD signal in the low frequency range of 0.01–0.10⍰Hz and divided it by the power of BOLD signal across the entire frequency range (0–0.25⍰Hz) to calculate fALFF^30^. Voxelwise fALFF was compared between responders and non-responders using t-tests implemented in SPM with age, sex, scanner, and movement (FD) as nuisance covariates, and applying a height threshold of p<0.005 and FDR-corrected cluster size p<.05.

## Results

### Baseline-biomarker analysis

Compared to non-responders, patients who would later meet strict criteria for clinical response demonstrated significantly greater baseline fALFF in bilateral orbitofrontal cortex (OFC, Figure 1). By contrast, no significant clusters were identified in which non-responders showed greater baseline fALFF compared to responders.

**Figure 1.**
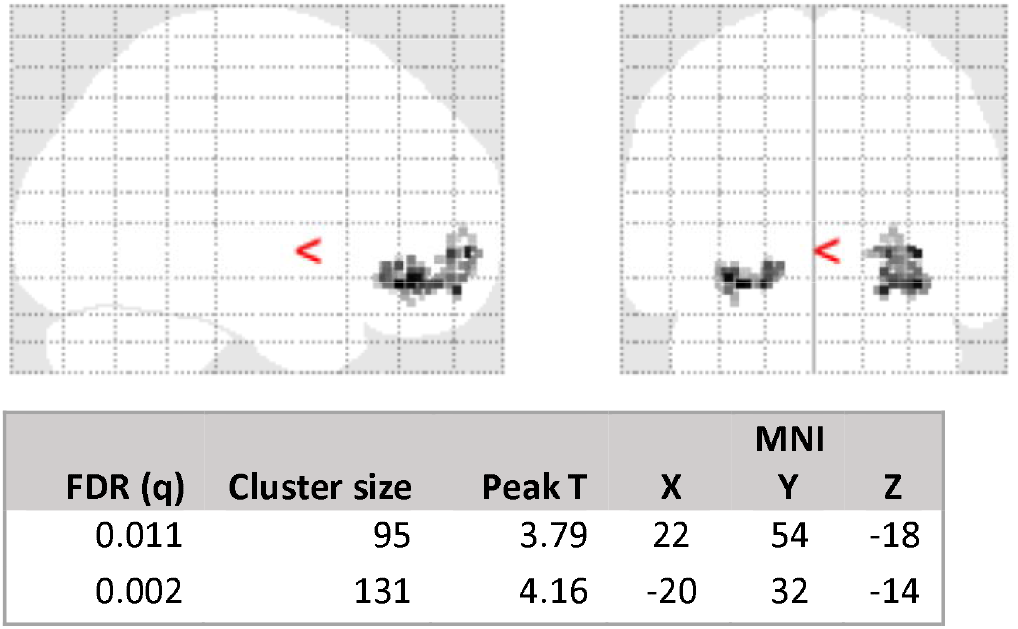

### Change-biomarker analysis

Responders (n=41) demonstrated significant increases in fALFF, from baseline to follow-up, in bilateral dorsal prefrontal cortex (middle/superior frontal gyrus) and retrosplenial cortex (posterior cingulate/precuneus; Figure 2a). Non-responders, by contrast, demonstrated significant fALFF increases in right premotor cortex / supplementary motor region (Figure 2b). No regions demonstrated significant decreases in fALFF over time in either group.

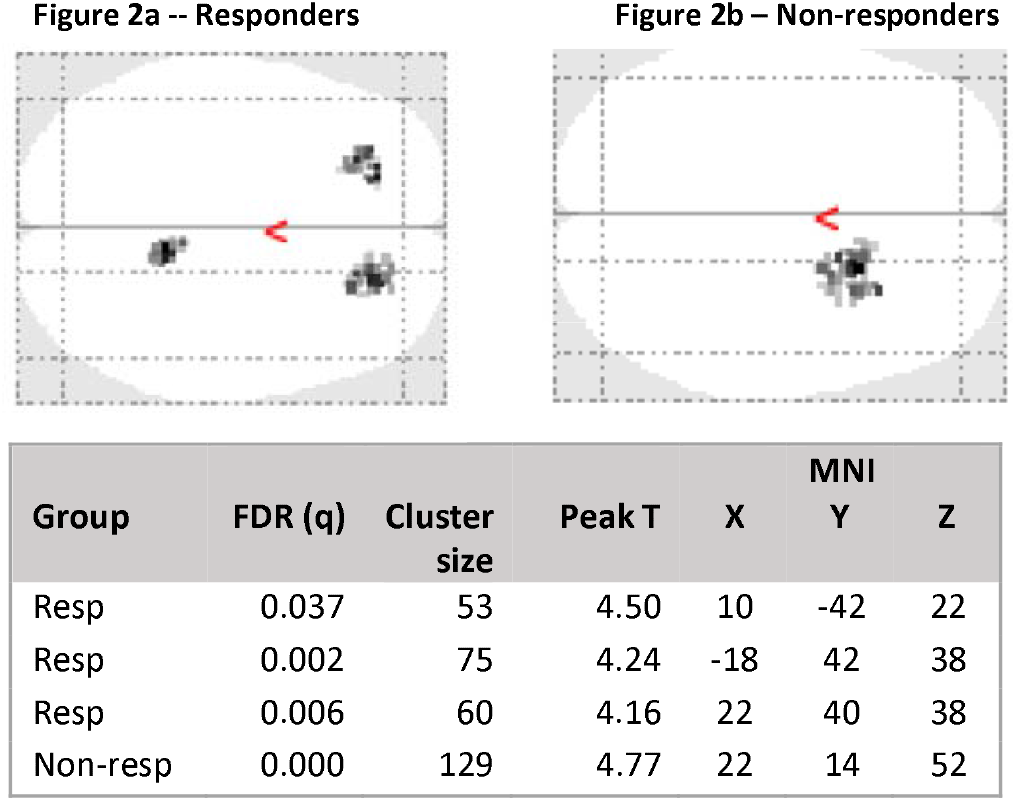

To determine if any of the changes observed in the patient groups were a function of normal test-retest variation, we examined change in fALFF in n=45 healthy comparison subjects scanned at two timepoints, 12 weeks apart. Seven brain regions demonstrated significant increases in fALFF from baseline to followup (encompassing bilateral sensorimotor cortex, bilateral lingual gyrus, left premotor cortex, left supplementary motor area, and right superior occipital cortex; see Figure 3). Notably, all of these regions were centered in posterior cortex (no peak or subpeak had y coordinate > 0), whereas both patient groups demonstrated fALFF increases in frontal regions only.

**Figure 3.**
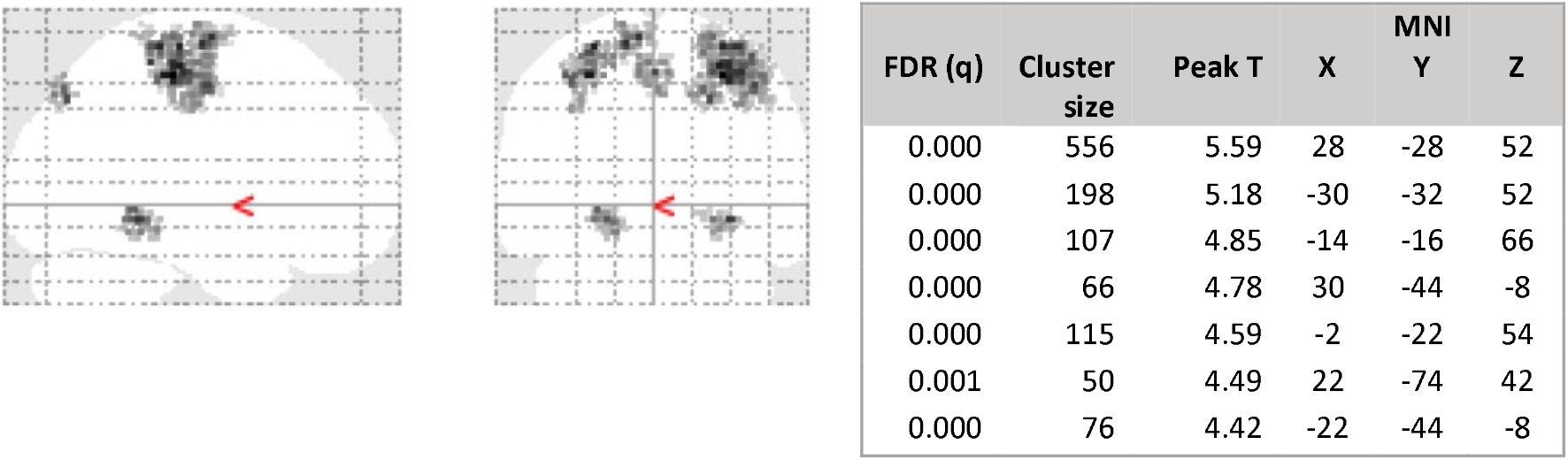

## Discussion

In the largest prospective study to date of resting state fMRI in first-episode psychosis, we report that activity in several prefrontal regions at rest may serve as treatment-relevant biomarkers. First, we observed that fALFF in bilateral orbitofrontal cortex at baseline may serve as a prognostic biomarker. Specifically, patients who later demonstrated robust positive symptom response to risperidone or aripiprazole had greater orbitofrontal fALFF at baseline, compared to patients who failed to respond in their initial 12-week trial of these antipsychotic agents. This result complements our previous finding that baseline striatal connectivity to multiple cortical regions (including OFC) may also have value as a baseline prognostic biomarker^21^. While our prior study, like most of the baseline-biomarker studies summarized in Table 1, employed functional connectivity analysis based on a hypothesis-driven striatal region-of-interest, the current study was not constrained by prior hypotheses. To the extent that fALFF indexes underlying neural activity^31-34^, our results suggest that first episode patients with reduced OFC activity are less likely to respond to conventional dopamine D2 agents. While a variety of clinical and preclinical studies have implicated OFC dysfunction in schizophrenia^16,61,62^, potentially indexing abnormal reward prediction^63^ and/or reversal learning^64^, the present study suggests that these abnormalities may especially be marked in treatment non-responsive patients. Further studies, in larger sample sizes, will be required to determine the potential sensitivity and specificity of OFC fALFF as a prognostic biomarker. It is also noteworthy that activity and connectivity of the OFC is especially sensitive to dopaminergic modulation^65^, but similar effects may also be elicited by modulation of the metabotropic glutamate receptor^66,67^, suggestive a possible target for future treatment development.

While fALFF in the ventral prefrontal cortex may serve as a baseline biomarker, results of the present study suggest that fALFF in dorsal prefrontal cortex may serve as a change biomarker. Increased fALFF in dorsal PFC over time was seen in subjects responding to treatment. These results replicate, in part, findings initially reported by Lui et al.^42^ which have since not been replicated by several small, uncontrolled studies^43,56,59^. Moreover, these results complement our previous findings that increases in dorsal prefrontal connectivity to the striatum (specifically, the dorsal caudate) were correlated with symptom improvement in a partially overlapping cohort of first episode patients^60^. While abnormalities of the dorsal prefrontal cortex have been a major focus of schizophrenia research for decades^68^, its role in treatment response has only recently come into focus. Several task-based fMRI studies have suggested that normalization of hypoactivity in the dorsal prefrontal cortex may be a marker of antipsychotic efficacy^69^, consistent with results of cross-sectional studies comparing treatment-resistant to treatment-responsive patients with schizophrenia^70^. A mechanistic understanding of dorsal prefrontal cortex suggests that tight regulation of calcium-cAMP signaling is required for optimal function, and that several pharmacologic approaches may be tested that can ameliorate calcium dysregulation^71^. Intriguingly, emerging evidence suggests that non-pharmacologic approaches to schizophrenia treatment such as repetitive transcranial magnetic stimulation (rTMS) and cognitive remediation therapy may also effectively modulate activity and connectivity in the dorsal prefrontal cortex^72^.

Additionally, we observed increased fALFF in the precuneus/posterior cingulate over the course of treatment in responders; this change was not observed in non-responders. This finding complements two recent studies reporting that increased connectivity of the posterior cingulate was associated with clinical response to antipsychotic treatment in the first episode of illness^53,54^. This brain region is considered the hub of the default mode network, and is critical to the ability to shift between self-oriented processing and (external) task-oriented function mediated by dorsal prefrontal cortex^73,74^. Collectively, our findings are consistent with a recently proposed model of schizophrenia^61^ that places the inability to shift between internal and external focus at the core of the disorder, and suggests novel pharmacologic and molecular approaches to ameliorating these deficits.

## Data Availability

Portions of the dataset reported herein have been uploaded to the NIMH data archive (Project #2475).

## Acknowledgements

This work was supported by grants from the National Institutes of Health: R01 MH108654 (PI: Malhotra); P50 MH080713 (PI: Malhotra); and R21 MH101746 (PIs: Robinson and Szeszko).

